# Validation of Generative AI Techniques for Synthetic Data Generation in Multiple Sclerosis Research: A Comparison with Real-World Evidence from the Italian MS Registry

**DOI:** 10.1101/2025.11.13.25340076

**Authors:** Pietro Iaffaldano, Saverio D’Amico, Giuseppe Lucisano, Massimiliano Copetti, Tommaso Guerra, Maria Assunta Rocca, Francesco Patti, Giovanna De Luca, Diana Ferraro, Rocco Totaro, Vincenzo Brescia Morra, Giuseppe Salemi, Emilio Portaccio, Matteo Foschi, Matilde Inglese, Maria Gabriella Coniglio, Clara Grazia Chisari, Francesca Caputo, Damiano Paolicelli, Mario Alberto Battaglia, Matteo Giovanni Della Porta, Victor Savevski, Mattia Delleani, Filippo Emanuele Colella, Elisabetta Sauta, Maria Pia Amato, Massimo Filippi, Maria Trojano, the Italian Multiple Sclerosis Register

## Abstract

**Importance:** Large multiple sclerosis (MS) registries provide crucial real-world evidence but often suffer from missing data, inconsistencies, and privacy limitations that restrict data sharing. The use of generative AI to create synthetic data (SD) is an emerging strategy to enhance real-world evidence research potentially overcoming these challenges.

**Objective:** To evaluate the validity of AI-generated synthetic data (SD) in replicating real data collected in the Italian MS and Related Disorders Register (RISM), and to compare the risk of progression independent of relapse activity (PIRA) between early intensive treatment (EIT) versus escalation treatment strategy (ESC) in both real and synthetic MS cohorts.

**Design, Setting, and Participants:** This validation study analyzed data from RISM. AI-based generative models were trained on a sub-cohort of 1,666 patients with tabularized MRI data to generate a synthetic dataset of 4,878 patients. SD was evaluated using the Synthetic vAlidation FramEwork powered by Train (SAFE), assessing fidelity, utility, and privacy. Clinical Synthetic Fidelity (CSF) and Nearest Neighbor Distance Ratio (NNDR) were used for statistical and privacy validation. Treatment outcome comparisons between EIT and ESC strategies were conducted for clinical validation using both real and synthetic datasets, focusing on the risk of PIRA.

**Exposures:** Initial disease-modifying therapy strategy, categorized as EIT versus ESC.

**Main Outcomes and Measures:** Primary outcome was the occurrence of PIRA, defined as confirmed disability accrual independent of relapses. Validation metrics included Clinical Synthetic Fidelity (CSF ≥90 optimal) and Nearest Neighbor Distance Ratio (NNDR, range 0.60–0.85 for privacy).

**Results:** The synthetic dataset demonstrated high fidelity (CSF=97%) and privacy preservation (NNDR=0.61). Treatment effect estimates for ESCs vs EIT were consistent across real and synthetic datasets, with largely comparable trends, with increased statistical significance in SD. Cox proportional hazards models confirmed the robustness of synthetic data in estimating the risk of the first PIRA event.

**Conclusions and Relevance:** AI-generated synthetic data reliably replicated treatment effect outcomes from real-world RISM data, overcoming missing data and providing a privacy-preserving alternative for data sharing and clinical research.

**Key points:** *Question:* Can Artificial Intelligence (AI)-generated synthetic data (SD) reliably replicate multiple sclerosis (MS) registry data and provide robust insights into progression independent of relapse activity (PIRA) phenomena under different treatment strategies?

*Findings:* In a cohort of 4,878 relapsing-onset MS patients from the Italian MS Register, AI-generated SD achieved high fidelity (CSF = 97%), and reproduced treatment effect outcomes. Both real and synthetic cohorts consistently showed that early intensive therapy reduced the risk of PIRA compared with an escalation strategy.

*Meaning:* SD can complement and enhance registry-based research by addressing missing data and supporting reproducible analyses in MS.

## 1. Introduction

Neuroinflammation and neurodegeneration phenomena define the course of multiple sclerosis (MS). (1–2) From the earliest stages of the disease and throughout its progression, irreversible neurological impairment accumulates as a result of both relapse-related worsening (RAW) and progression independent of relapse activity (PIRA) events. (3–5) PIRA is becoming more widely acknowledged as the primary cause of disability progression: understanding how disease modifying therapies (DMTs) can impact PIRA events is crucial to unraveling the intricate pathways of silent progression and improving treatment approaches in MS. (6) Real-world evidence (RWE) from large MS registries provided significant insights into disease progression in recent years. (7) However, real-world datasets frequently are plagued by missing information and discrepancies, and data sharing between registries and other data sources are often restricted due to data privacy issues. Generative artificial intelligence (AI) offers a promising avenue to overcome such issues by producing synthetic data (SD), trained to learn the key features of a real source dataset, and offers a potential solution to circumvent some of these challenges. (8–10) The application of AI-driven SD in various research fields, such as hematology, oncology and neurology has shown encouraging results, demonstrating that SD can improve methodologies while preserving high fidelity to real-world distributions (11–13) and can accelerate research focused on personalized precision medicine. In this study, we aimed to: 1) apply innovative synthetic data generation methods to the real data collected in the Italian MS and Related Disorders register (RISM); 2) implement a validation framework to evaluate synthetic data fidelity and privacy preservability and 3) to evaluate the validity of AI-generated SD to replicate, and to compare the risk of PIRA between early intensive treatment (EIT) versus escalation treatment strategy (ESC) in both real and synthetic MS cohorts.

## 2. Methods

### 2.1 Data Source and Study Population

This was a study based on data extracted from the RISM database. RISM was approved by the ethical committee at the “Azienda Ospedaliero – Universitaria – Policlinico of Bari” (Study REGISTRO SM001 – approved on 8 July 2016) and by local ethics committees in all participating centers. Patients signed an informed consent allowing the collection and use of their clinical data for research purposes. According to the Registry rules, the Scientific Committee of the RISM register approved to conduct this project and extract and use the registry data. Data extraction for this study was executed in March 2023. The selection criterias for patients included relapsing-onset MS patients with a minimum follow-up of five years, an initial DMT prescription within three years of disease onset, and at least three Expanded Disability Score (EDSS) evaluations following treatment initiation. Patient data retrieved from the register included demographic, clinical, magnetic resonance imaging (MRI) and treatment-related characteristics.

### 2.2 Synthetic Data Generation and Validation

Generative Adversarial Networks (GAN) and Tabular Generative Pretrained Transformer (T-GPT) were implemented as generative models and were trained in a sub-cohort of patients with complete MRI tabular information to generate synthetic data of MS patients. MRI information refers to tabular variables extracted from clinical practice records and entered into the RISM. The T-GPT was chosen due to better performances and for the nature of clinical data. The generative model was trained on patients with complete data to generate new synthetic cohorts with complete information. Full architectures, preprocessing, hyperparameters, and training settings are provided in Supplementary Material S1 (SF3, SF4, SF5).

The synthetic cohort was evaluated using the Synthetic vAlidation FramEwork powered by Train (SAFE) (14), to assess statistical fidelity, clinical utility, and privacy preservability, ensuring that the generated data accurately reflected real-world distributions without compromising patient confidentiality. Statistical and clinical fidelity were measured using the Clinical Synthetic Fidelity (CSF) score, which averages multiple statistical tests; an optimal score is considered >90%. To assess privacy and the risk of a synthetic record matching an original patient, we first measured the identical match share (IMS) (exact matches between synthetic and original data). We also calculated the distance to the closest record (DCR), which is the absolute distance from a synthetic record to its nearest original counterpart. Furthermore, we used the Nearest Neighbor Distance Ratio (NNDR) as the ratio of the distances from a synthetic record to its nearest and second-nearest neighbors. The NNDR optimal range is 0.60-0.85, allowing for a fair comparison of inliers and outliers in the population (15). SAFE library specifications and detail are reported in Supplementary SF6).

### 2.3 Clinical Validation and Statistical Analysis

For clinical validation of synthetic data we replicated the risk of PIRA comparison analysis of EITs versus ESCs in real and synthetic MS cohorts. Patients were categorized into two EIT and ESC groups, based on the initial DMT prescribed. The EIT cohort included patients who initiated treatment with high efficacy (HE)-DMTs: natalizumab, alemtuzumab, ocrelizumab, cladribine, fingolimod, or mitoxantrone. Conversely, the ESC group included patients initially treated with moderate efficacy (ME)-DMTs, such as interferon-beta products, glatiramer acetate, teriflunomide, dimethyl fumarate, and escalated to HE-DMTs after at least one year. Confirmed disability accrual (CDA) was defined as a confirmed 6□month disability increase from study baseline, measured by EDSS (increase ≥1.5 points with baseline EDSS□=□0; ≥1.0 point with baseline EDSS >1.0, and□<5.5; ≥0.5 point with baseline EDSS >6.0).

The date of CDA was assigned at the first EDSS when an increase was registered. PIRA, the primary outcome, was defined as a CDA event occurring more than 90□days after and more than 30□days before the onset of a relapse. The baseline was represented by the first prescription of DMT.

In descriptive analyses, categorical data were expressed as frequency and proportion. Continuous data were expressed as median and interquartile range (IQR).

Clinical characteristics were compared and tested between EIT and ESC groups by chi-squared and T-test, for categorical and continuous variables respectively. For continuous variables, normal distribution assumption was checked by Shapiro-Wilk test and visual inspection of Q-Q plot.

For correlation analysis, the relationships between the columns of the dataset were evaluated by building correlation matrices. Since the clinical and demographic features had different data types (i.e. continuous and categorical), we used different methods to calculate the correlation metric. We calculated the correlation of continuous-continuous features using Pearson’s correlation. For continuous-categorical features, we used the Correlation Ratio and for categorical-categorical features we performed the Uncertainty Coefficient proposed by Theil’s U. We created two correlation matrices for real and synthetic datasets respectively, then we calculated the difference between those matrices. Metric was calculated as the average of all values in the difference matrix. Cox proportional hazards models were used to assess the risk of PIRA in EIT and ESC groups in both real and SD cohorts of 4,878 subjects. The following variables have been included as covariates in the models: gender (male vs. female), age and EDSS at first DMT start, time to first DMT start from onset, number of relapses in the two years before DMT start (0, 1 or ≥2), onset type (multifocal vs monofocal), T2 lesion number (0, 1-2, 3-8 or ≥9), gadolinium (GD)-enhancing T1 lesion (presence vs absence) at the MRI performed before DMT start, treatment strategy (ESC as reference).

The proportional hazards (PH) assumption was checked testing the logarithm of time interaction with covariates and by Schoenfeld residuals. Results of Cox regression models were expressed as hazard ratio (HR) and 95% confidence interval (95%CI) of reaching the outcomes. The homogeneity of HR across datasets was assessed to validate SD.

P-values <0.05 result in statistical significance. The software used to perform the analyses is R 4.2.3.

## 3. Results

Clinical data of 79,001 patients were available in the RISM at the time of data extraction. After applying the inclusion criteria, we retrieved a cohort of 4,878 RMS patients. Among them, 914 patients were treated with an EIT approach, while 3,964 patients followed an ESC strategy. Baseline characteristics, stratified on the basis of the treatment strategy, are shown in Table 1. The two groups differed significantly in several baseline characteristics. Patients in the EIT group were slightly older at the initiation of the first DMT compared to those in the ESC group (30.85 vs. 29.70 years, p=0.0018). The proportion of patients over 40 years old was higher in the EIT group (25.82% vs. 20.41%), whereas the proportion of younger patients (up to 20 years old) was lower (11.27% vs. 13.07%). No significant difference was found in gender distribution between the two groups (p=0.9549). Patients in the EIT group presented higher median EDSS scores compared to ESC group at the first DMT start (2.5 vs 1.5, respectively). At baseline, the lesion burden was also significantly higher for patients in the EIT group, with a larger proportion presenting with at least nine T2 lesions compared to the ESC group (21.44% vs 16.15%, respectively).

**Table 1.**
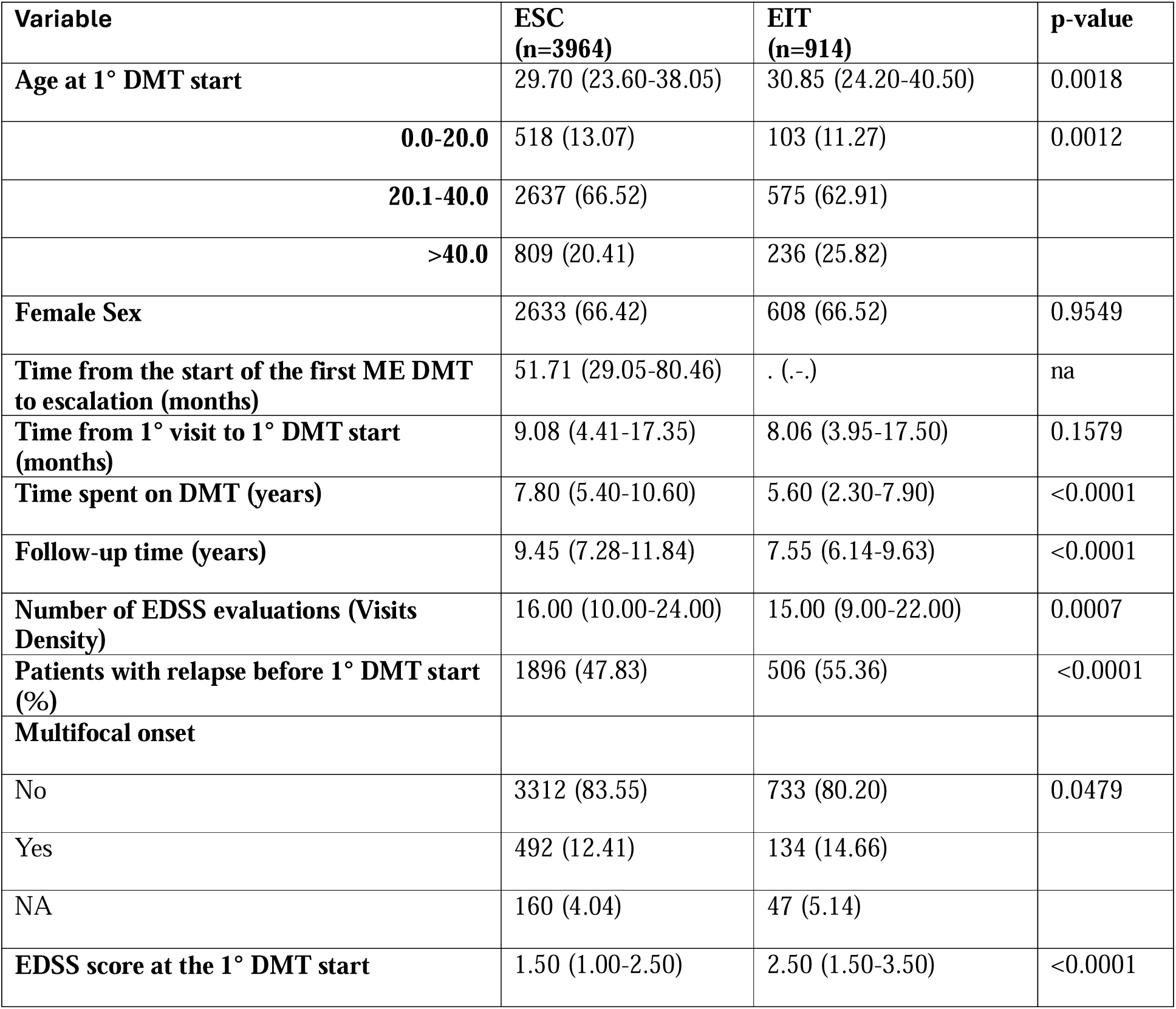
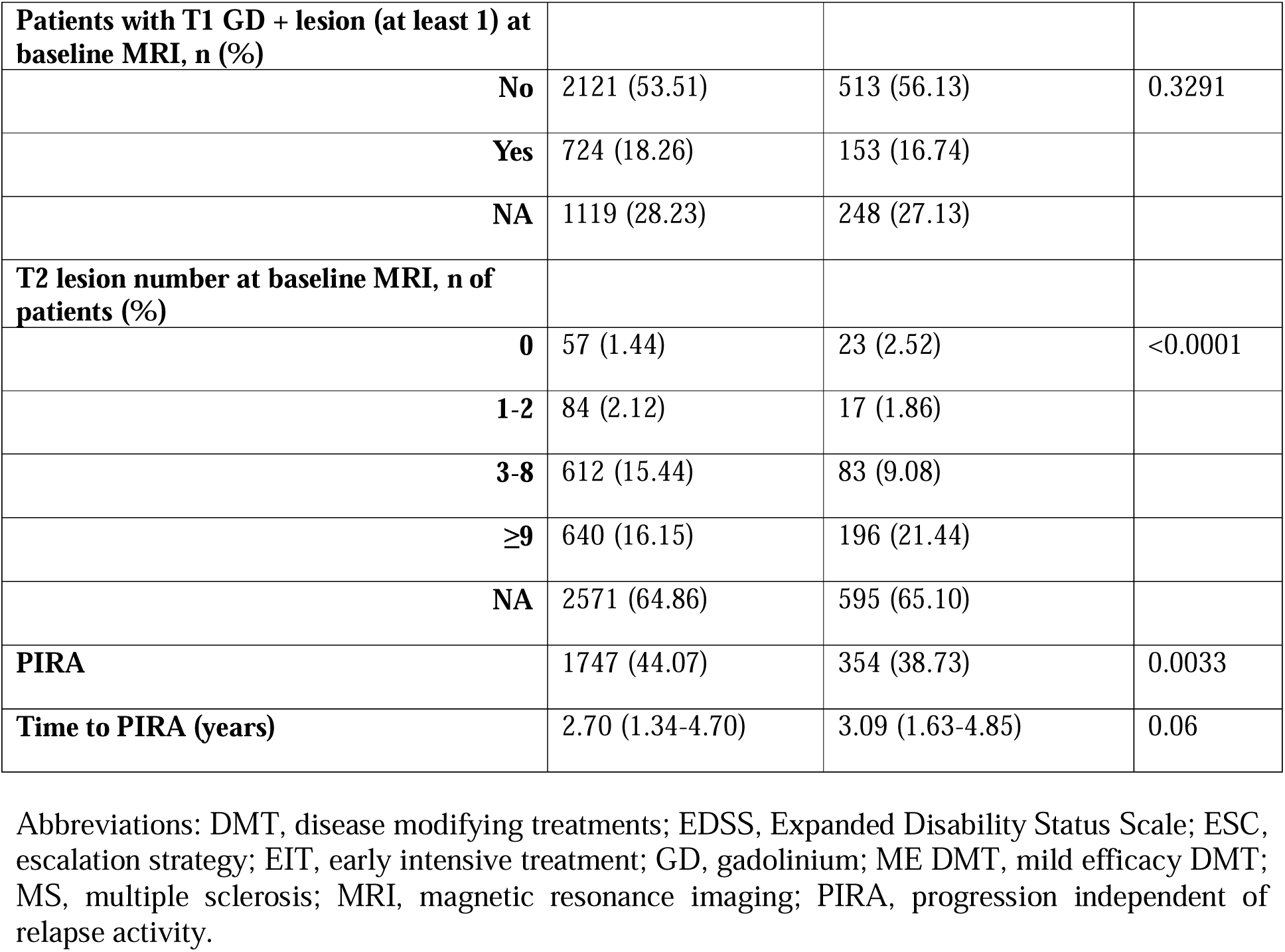
Baseline clinical and demographic characteristics of EIT and ESC cohort.

| **Variable** | **ESC**<br>**(n=3964)** | **EIT**<br>**(n=914)** | **p-value** |
| --- | --- | --- | --- |
| **Age at 1° DMT start** | 29.70 (23.60-38.05) | 30.85 (24.20-40.50) | 0.0018 |
| **0.0-20.0** | 518 (13.07) | 103 (11.27) | 0.0012 |
| **20.1-40.0** | 2637 (66.52) | 575 (62.91) |  |
| **>40.0** | 809 (20.41) | 236 (25.82) |  |
| **Female Sex** | 2633 (66.42) | 608 (66.52) | 0.9549 |
| **Time from the start of the first ME DMT to escalation (months)** | 51.71 (29.05-80.46) | . (-.) | na |
| **Time from 1° visit to 1° DMT start (months)** | 9.08 (4.41-17.35) | 8.06 (3.95-17.50) | 0.1579 |
| **Time spent on DMT (years)** | 7.80 (5.40-10.60) | 5.60 (2.30-7.90) | <0.0001 |
| **Follow-up time (years)** | 9.45 (7.28-11.84) | 7.55 (6.14-9.63) | <0.0001 |
| **Number of EDSS evaluations (Visits Density)** | 16.00 (10.00-24.00) | 15.00 (9.00-22.00) | 0.0007 |
| **Patients with relapse before 1° DMT start (%)** | 1896 (47.83) | 506 (55.36) | <0.0001 |
| **Multifocal onset** |  |  |  |
| No | 3312 (83.55) | 733 (80.20) | 0.0479 |
| Yes | 492 (12.41) | 134 (14.66) |  |
| NA | 160 (4.04) | 47 (5.14) |  |
| **EDSS score at the 1° DMT start** | 1.50 (1.00-2.50) | 2.50 (1.50-3.50) | <0.0001 |
| **Patients with T1 GD + lesion (at least 1) at baseline MRI, n (%)** |  |  |  |
| **No** | 2121 (53.51) | 513 (56.13) | 0.3291 |
| **Yes** | 724 (18.26) | 153 (16.74) |  |
| **NA** | 1119 (28.23) | 248 (27.13) |  |
| **T2 lesion number at baseline MRI, n of patients (%)** |  |  |  |
| **0** | 57 (1.44) | 23 (2.52) | <0.0001 |
| **1-2** | 84 (2.12) | 17 (1.86) |  |
| **3-8** | 612 (15.44) | 83 (9.08) |  |
| **≥9** | 640 (16.15) | 196 (21.44) |  |
| **NA** | 2571 (64.86) | 595 (65.10) |  |
| **PIRA** | 1747 (44.07) | 354 (38.73) | 0.0033 |
| **Time to PIRA (years)** | 2.70 (1.34-4.70) | 3.09 (1.63-4.85) | 0.06 |

Abbreviations: DMT, disease modifying treatments; EDSS, Expanded Disability Status Scale; ESC, escalation strategy; EIT, early intensive treatment; GD, gadolinium; ME DMT, mild efficacy DMT; MS, multiple sclerosis; MRI, magnetic resonance imaging; PIRA, progression independent of relapse activity.

A sub-cohort of 1,666 MS patients with non-missing MRI information (tabularized) was selected and was used to train a T-GPT to generate a synthetic dataset of 4,878 patients with complete data. The flowchart reporting patients’ selection procedure and cohorts’ definition is shown in Figure 1. SD demonstrated a strong resemblance to real distributions, with high alignment in variables such as age at DMT initiation, sex distribution, baseline EDSS, lesion load at baseline MRI, and time to first PIRA event. (Figure 2). SD closely mirrored real-distributions, with a CSF score of 97% confirming high fidelity. Privacy was preserved, with an NNDR value of 0.61 ensuring no identifiable patient data leakage. The robustness of the synthetic data cohort was also highlighted by the Spearman/Pearson/Kendall correlation matrices shown in Figure 3. Both strength and direction of correlation were very similar in SD and real data and the correlation differences between SD and real data were close to zero, as shown in Figure 3.

**Figure 1.**
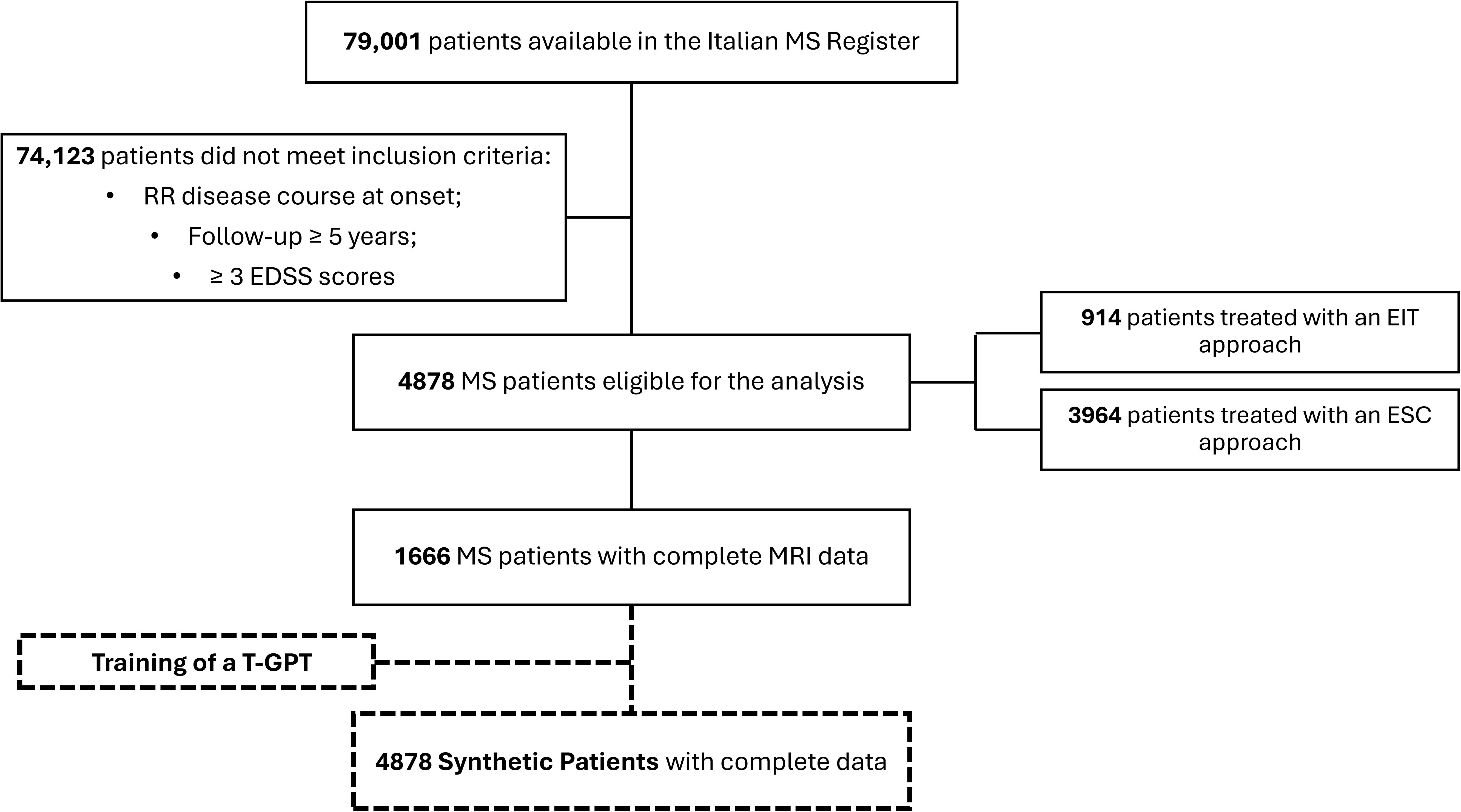
Flowchart of patient selection procedure. Abbreviations: early intensive treatment, EIT; escalation treatment strategy, ESC; Expanded Disability Status Scale, EDSS; magnetic resonance imaging, MRI; relapsing remitting multiple sclerosis, RRMS; Tabular Generative Pretrained Transformer, T-GPT;

**Figure 2.**
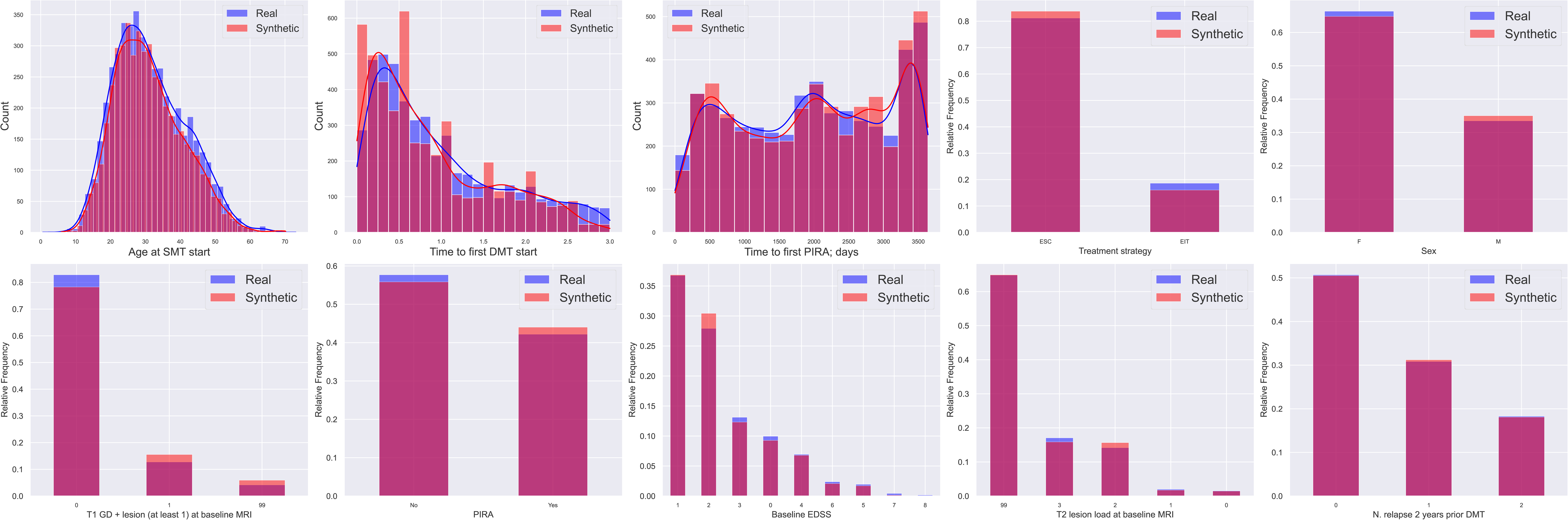
Comparison of distributions for key clinical variables between real and synthetic datasets. Distributions of key demographic and clinical variables are shown for both real and synthetic data. Variables include: age at start of disease-modifying therapy (DMT), sex, baseline Expanded Disability Status Scale (EDSS), treatment strategy (ESC vs EIT), presence of T1 gadolinium-enhancing lesions at baseline MRI, T2 lesion load at baseline, number of relapses in the two years prior to DMT initiation, time to first DMT initiation, and time to first progression independent of relapse activity (PIRA) event. Synthetic data closely mirror the real data across all distributions, supporting the fidelity of the generative model.

**Figure 3.**
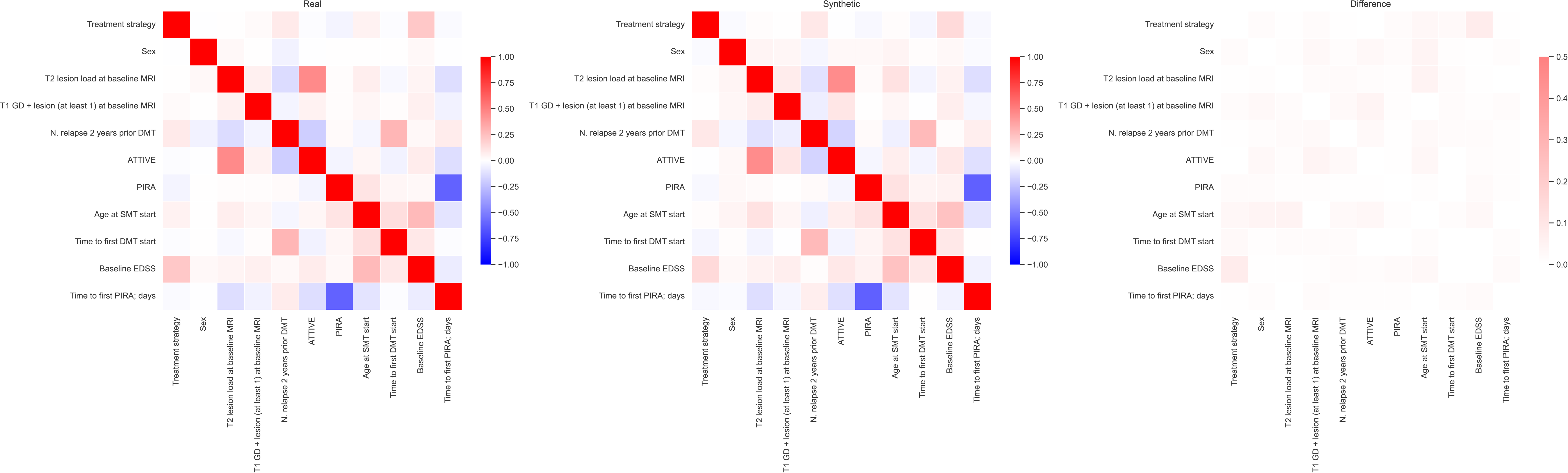
Correlation comparison between the real and synthetic data.

Cox proportional hazards models were used to assess the impact of treatment strategy on PIRA risk in the real cohort and SD group. The analyses included all covariates reported in the Methods section. The age at DMT start emerged as a risk factor of PIRA (HR, 95% CI, 1.02, 1.01–1.03, *p*<0.001 for the real data; 1.02, 1.01–1.02, *p*<0.001 for SD). The models retrieved overlapping results, but an increase in the statistical significance of the associations was observed when the Cox models were applied to SD. The EIT approach in reducing the risk of PIRA compared to ESC was not significant in the real cohort (HR: 0.88; 95% CI: 0.71-1.08, *p*=0.22), and significant in the SD (0.86, 0.76-0.98, *p*=0.02). A longer time to the initiation of the first DMT resulted also a significant risk factor for PIRA in the SD (HR: 1.10, 95% CI: 1.04-1.17, *p<0.001).* The effect of MRI variables on PIRA risk retrieved significative findings. Patients with ≥9 T2 lesions at baseline had a significantly higher risk of PIRA (HR=1.88, 95% CI: 1.20-2.96, p=0.01), and conversely the presence of GD-enhancing T1 lesion (HR=0.82, 95% CI: 0.73-0.91, *p*<0.001) was a protective factor of PIRA events. Conversely, MRI parameters were not significant in the real cohort group. Forest plots reporting HR in real and synthetic datasets are shown in Figure 4. Exact HR with CIs are provided for both real and synthetic datasets in Supplementary Material (SF7).

**Figure 4.**
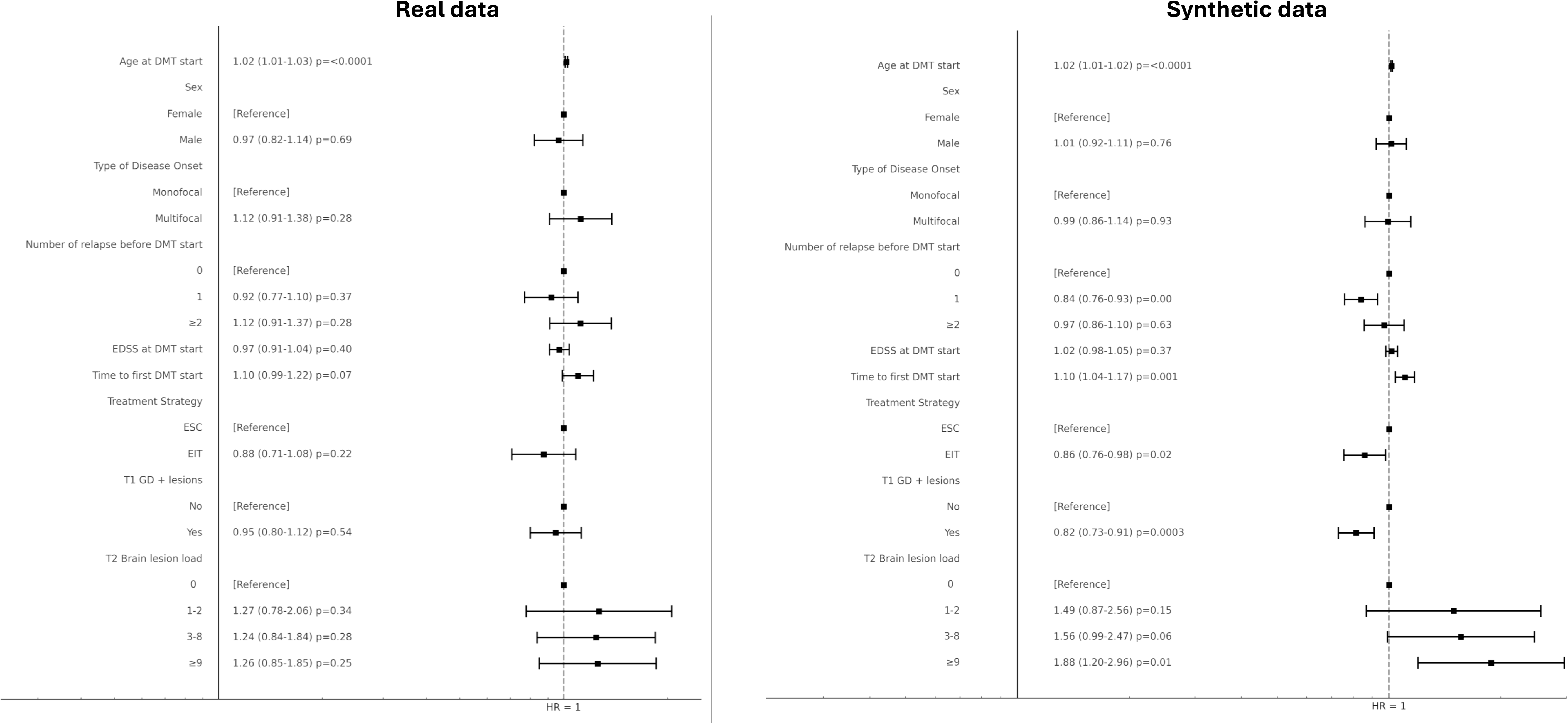
Forest Plots for Cox proportional hazards model applied to the real and synthetic datasets to investigate risk factors of PIRA.

## 4. Discussion

Our findings support the validity of AI-generated SD to replicate clinical and MRI real data as those collected in the RISM with high fidelity (CSF=97%) and privacy preservation (NNDR=0.61) values. Recent literature highlights the increasing role of SD in various fields of biomedical research, particularly in enhancing study reproducibility and overcoming data-sharing restrictions (8, 12, 13, 16–18). Research on neurological diseases has also been greatly impacted by recent developments in generative AI modeling, which have opened up novel possibilities for data augmentation, privacy protection, and personalized medicine. Variational autoencoders and GANs have proven to be effective in producing high-fidelity synthetic neuroimaging data, which has been successfully applied in neurodegenerative diseases. (19, 20) Synthetic data can also facilitate external validation of predictive models, providing robust insights into treatment effects. (21) For research purposes, synthetic datasets that replicate a portion of the Public Health England Cancer Registry and the Netherlands Cancer Registry are now accessible in Europe (22, 23) and the USA (24).

We demonstrated that using the synthetic MS cohort, the outcomes of the comparative effectiveness analysis between an ESC and an EIT strategy on the risk of the first PIRA occurrence were consistent with those obtained using the real MS cohort. The Cox model confirmed that EIT strategy significantly reduced the risk of PIRA. Several previous observational RWE from large cohorts reported significant reductions in long-term disability with EIT compared to ESC paradigms. (25, 26) Another study from the I-MS&RD research group assessed long-term disability trajectories and demonstrated that the mean yearly delta-EDSS values were significantly higher following ESC approaches, emphasizing the therapeutic effects of early initiation of HE DMTs (27). Our results, consistent across both real and synthetic cohorts, reinforced these findings.

Most importantly, this study highlights the potential of SD to complement real world data. Indeed, the use of AI-generated SD enabled a more comprehensive evaluation of MRI-derived risk factors for disability progression. Previous work using register data has often not allowed the use of MRI data, because frequently missed due to the lack of a systematic acquisition of these data. (28) The synthetic dataset not only confirmed the previously reported association between high lesion load and increased PIRA risk (29, 30) but also provided an additional layer of validation by preserving these relationships in an independent dataset. Same consideration applied to the presence of inflammatory activity on MRI, evidenced by the detection of GD-enhancing T1 lesion, which resulted conversely a protective factor of PIRA occurrence. Therefore, our findings suggest that SD could constitute a valuable tool for validating MRI-based prognostic models, offering a novel framework to enhance the robustness of real-world evidence in MS. The ability of SD to mimic the effects of real-world therapy on PIRA has become primarily important as PIRA is increasingly acknowledged as a significant contributor to disability accrual in MS. (1–6, 31, 32)

One of the most promising applications of generative modeling in neurological disease research suggested by this study is its potential to contribute to precision medicine by creating digital twins of patients. In MS research, digital twin approaches based on SD could enable real-time decision-making for clinicians, helping to tailor treatment strategies to individual patient profiles while maintaining data privacy. (19)

While the advantages of generative AI in medical research are substantial, challenges remain, and limitations must be acknowledged. Subtle biases in the original dataset may be perpetuated, and the quality of the real-world training data determines how well SD is generated. Model evaluation, data fidelity, and privacy concerns must be rigorously addressed clinically relevance and high ethical standards preservation. Additionally, further validation is needed to assess its applicability across different MS populations and treatment paradigms.

## 5. Conclusion

In conclusion, this study confirms that the use of SD is a reliable method for complementing RWE reducing the burden of missing data such as MRI-derived data. Future research should explore the broader application of SD in MS research, including its potential use in multicenter studies and regulatory decision-making and refining SD methodologies to maximize their impact in neurological disease research.

## Data Availability

Anonymized data will be available upon reasonable request to the authors of a qualified investigator.

## Author Contributions

PI and MT had full access to all the data in the study and took responsibility for the integrity of the data and the accuracy of the data analysis. Concept, design and statistical analysis: PI, SDA, GL, MC, TG, MT. All authors contributed to acquisition, analysis, interpretation of data and to the drafting of the manuscript.

## Data sharing

Anonymized data will be shared on reasonable request from a qualified investigator.

## Acknowledgments

The authors thank the Research Assistants of the RISM register.

