## Supplementary material for "Validation of Generative AI Techniques for Synthetic Data Generation in Multiple Sclerosis Research: A Comparison with Real-World Evidence from the Italian MS Registry"


SUPPLEMENTARY_FILE_1 (SF1) – Glossary of Abbreviations 2
1 Clinical Terms, Organizations and Affiliations 2
2 Statistical and Evaluation Metrics 5
3 Data types, Models and Frameworks 7
SUPPLEMENTARY_FILE_2 (SF2) – Study population 7
SUPPLEMENTARY_FILE_3 (SF3) – Generative models for Synthetic Data 8
1 GANs 8
2 T-GPT 9
SUPPLEMENTARY_FILE_4 (SF4) – Data pre-processing 9
SUPPLEMENTARY_FILE_5 (SF5) – Training strategies and implementation details 10
SUPPLEMENTARY_FILE_6 (SF6) – Development of SAFE 12
1 Fidelity metric 12
2 Correlation analysis 13
3 Privacy metric 14
4 Clinical validation 14
SUPPLEMENTARY_FILE_7 (SF7) – Experimental results settings 15
1 Fidelity 15
2 Correlation 16
3 Privacy 16
4 Clinical validation and utility 17
References 19

**SUPPLEMENTARY_FILE_1 (SF1) – Glossary of Abbreviations**

This glossary provides a comprehensive list of abbreviations used throughout the manuscript or the document, organized into three main categories: (1) Clinical Terms, Organizations and Affiliations (2) Statistical and Evaluation Metrics, and (3) Data Types, Models and Frameworks. It is intended as a quick reference to ensure clarity and consistency for readers across a range of interdisciplinary topics.

*1 Clinical Terms, Organizations and Affiliations*

| **Abbreviation** | **Full Name** | **Description** |
|---|---|---|
| AOU | Azienda Ospedaliero-Universitaria | Italian university hospital trust (hospital company). |
| ASM | Azienda Sanitaria Locale di Matera | Local health authority of Matera (hospital network). |
| CDA | Confirmed Disability Accrual | EDSS-based confirmed 6-month disability increase used to define progression events. |
| CORESEARCH | Center for Outcomes Research and Clinical Epidemiology (CORESEARCH) Srl | Italian CRO named in affiliations. |
| DINOGMI | Dipartimento di Neuroscienze, Riabilitazione, Oftalmologia, Genetica e Scienze Materno-Infantili | University of Genoa department. |
| DMT | Disease-Modifying Therapy | MS pharmacologic treatments; exposure variable and model covariate. |
| EDSS | Expanded Disability Status Scale | Ordinal disability scale in MS used for outcomes and baseline severity. |
| EIT | Early Intensive Treatment | Treatment strategy initiating with high-efficacy DMTs. |

| | | |
|---|---|---|
| ESC | Escalation Treatment Strategy | Treatment strategy starting with moderate-efficacy DMTs and escalating to high-efficacy DMTs. |
| FISM | Fondazione Italiana Sclerosi Multipla | Italian Multiple Sclerosis Foundation. |
| GD | Gadolinium | Contrast agent used to identify enhancing T1 lesions on MRI. |
| HE | High-Efficacy (DMTs) | Label for more potent disease-modifying therapies. |
| HE-DMTs | High-Efficacy Disease-Modifying Therapies | DMTs such as natalizumab, ocrelizumab, etc., used in the EIT strategy. |
| I-MS&RD | Italian Multiple Sclerosis & Related Disorders | Name component of the national register (basis for RISM). |
| IKNL | Integraal Kankercentrum Nederland | Netherlands Comprehensive Cancer Organisation (owner of the NCR). |
| IRCCS | Istituto di Ricovero e Cura a Carattere Scientifico | Italian designation for research hospitals. |
| MOSAIC | MOSAIC Project | Study/initiative cited in references. |
| MRI | Magnetic Resonance Imaging | Neuroimaging modality; baseline MRI variables included (T1 GD+ and T2 lesions). |
| MS | Multiple Sclerosis | Disease studied; basis of the RISM registry. |
| NCR | Netherlands Cancer Registry | National cancer registry (referenced in methods/discussion). |
| NEUROFARBA | Neurosciences, Psychology, Drug Research and Child Health | Univ. of Florence department (NEUROFARBA). |

| | | |
|---|---|---|
| PIRA | Progression Independent of Relapse Activity | A confirmed increase in disability that occurs outside relapse periods (relapse-independent). |
| RAW | Relapse-Related Worsening | Disability accrual associated with relapses. |
| RISM | Italian MS & Related Disorders Register | Nationwide registry providing the study dataset. |
| RMS | Relapsing Multiple Sclerosis | Clinical course comprising relapsing disease; subset analyzed. |
| RRMS | Relapsing-Remitting Multiple Sclerosis | Common MS relapsing phenotype. |
| SM001 | Study ID SM001 | Internal study identifier mentioned in manuscript. |
| SRL | Società a responsabilità limitata | Italian "limited liability company" legal form. |
| SS | San Salvatore / 'SS Annunziata' (contextual hospital names) | Appears in affiliation lines as part of hospital names. |
| T1 | T1-weighted MRI | MRI sequence; T1 GD-enhancing lesions are a predictor. |
| T2 | T2-weighted MRI | MRI sequence; T2 lesion counts included as covariates. |
| UOC | Unità Operativa Complessa | Complex operating unit (hospital department). |
| UOS | Unità Operativa Semplice | Simple operating unit (hospital sub-department). |
| USL | Unità Sanitaria Locale | Local health unit (regional healthcare provider). |

Table 1: List of clinical terms, organizations and affiliations abbreviations, full names and descriptions.

*2 Statistical and Evaluation Metrics*

| **Abbreviation** | **Full Name** | **Description** |
|---|---|---|
| - | Chi-squared test | |
| CI | Confidence Interval | A range of values that likely contains the true value of a population parameter<br>with a given confidence level (e.g., 95%). |
| - | Correlation Ratio | Measures how strongly a categorical variable explains variation in a continuous variable; it’s the proportion of between-group variance to total variance (0–1) and is asymmetric. |
| - | Cox proportional hazards models | Semi-parametric survival models estimating the effect of covariates on event risk. |
| CSF | Clinical Synthetic Fidelity | Metric introduced in the SAFE library quantifying similarity between clinical distributions in real vs synthetic data. |
| HR | Hazard Ratio | A metric quantifying how often a particular event occurs in one group compared to another group over time. It is the ratio of the hazard rates for two groups, representing the instantaneous risk of an event happening at any given point in time. |
| IQR | Interquartile Range | Summary statistic for continuous variables. |
| NNDR | Nearest Neighbor Distance Ratio | A metric quantifying the ratio of each synthetic record's distance to the nearest training neighbor compared to the distance to the second nearest training neighbor. It works as a privacy metric in synthetic data evaluation |
| r/τ/ρ | Pearson/Kendall/Spearman correlation | Linear correlation for continuous variables/Rank-based measure from concordant/discordant pairs/Rank-based correlation that captures monotonic relationships using ranks |
| PH | Proportional Hazards | Assumption/model framework for Cox regression. |

| | | |
|---|---|---|
| Q-Q | Quantile–Quantile | Plot used to assess normality in continuous variables. |
| - | Schoenfeld residuals | For Cox models, residuals at each event time equal to the observed covariate value minus its risk-set average. Used to test PH assumption. |
| - | Shapiro–Wilk test | Statistical test of normality for a sample. |
| - | T-test | Statistical test used to compare means between groups. |
| - | Uncertainty Coefficient (Theil's U) | Information-theoretic, directional association for categorical variables: proportion of uncertainty in Y reduced by knowing X. |

Table 2: List of statistical and evaluation metric abbreviations, full names and descriptions.

*3 Data types, Models and Frameworks*

| **Abbreviation** | **Full Name** | **Description** |
|---|---|---|
| AI | Artificial Intelligence | Computational methods; here used for generative modeling of synthetic data. |
| GAN | Generative Adversarial Networks | A deep learning model that generates synthetic data by training two neural<br>networks in competition. |
| RWE | Real-World Evidence | Observational evidence from registries like RISM. |
| SAFE | Synthetic vAlidation FramEwork powered by Train | A framework powered by Train [1], designed to validate the quality of synthetic<br>data. It assesses the fidelity, utility, and privacy of synthetic datasets, ensuring<br>they accurately represent real data while protecting sensitive information. |
| SD | Synthetic Data | Artificially generated data that mimics the statistical properties of real data,<br>used to enable research and testing while protecting patient privacy. |
| T-GPT | Tabular Generative Pretrained Transformer | A Transformer-based generative model tailored to work and generate tabular clinical data. |

Table 3: List of data type, model, framework, and platform abbreviations, full names and descriptions.

### SUPPLEMENTARY_FILE_2 (SF2) – Study population

This investigation utilized clinical data obtained from the Italian Multiple Sclerosis Register (RISM), a comprehensive repository of patient information maintained across multiple medical centers. The registry itself received formal ethical approval from the ethics committee of the Azienda Ospedaliero-Universitaria Policlinico of Bari under study protocol REGISTRO SM001, which was granted on July 8, 2016. Subsequently, ethics committees at each of the participating institutions provided their respective approvals, ensuring compliance with local regulatory requirements and ethical standards. All patients enrolled in the registry provided written informed consent that explicitly authorized the collection, storage, and utilization of their clinical information for research activities. For the present analysis, the Scientific Committee overseeing the RISM register reviewed and approved the research proposal, granting permission to access and analyze the relevant registry data.

The data extraction process for this investigation was completed in March 2023. The study population was defined according to specific inclusion criteria designed to ensure adequate longitudinal follow-up and standardized treatment approaches. Eligible participants were required to have a diagnosis of relapsing-onset multiple sclerosis with documented follow-up extending at least five years from their initial presentation. Additionally, patients must have started disease-modifying therapy within three years of their disease onset, reflecting early intervention strategies. To enable robust disability progression analyses, a minimum of three Expanded Disability Status Scale (EDSS) assessments recorded after treatment initiation was required for inclusion. The dataset extracted from the registry encompassed multiple dimensions of patient information, including demographic characteristics, comprehensive clinical assessments, magnetic resonance imaging findings in tabular format, and detailed treatment histories, thereby providing a multifaceted view of the disease course and therapeutic interventions.

### SUPPLEMENTARY_FILE_3 (SF3) – Generative models for Synthetic Data

To model the mixed, high-dimensional clinical variables extracted from the Italian MS Register (RISM), we implemented two complementary generative families: Generative Adversarial Networks (GANs) and a Transformer-based Tabular Generative Pretrained Transformer (T-GPT). Both models were trained on a sub-cohort of 1,666 patients with complete tabular MRI variables to synthesize a patient-level dataset free of missingness. From this sub-cohort, the T-GPT was fine-tuned and then used, together with the GAN baseline, to generate a synthetic cohort of 4,878 patients, matching the analysis cohort size; the choice of T-GPT reflected better performance for the clinical tabular setting reported in our manuscript.

#### *1 GANs*

Generative Adversarial Networks (GANs), first introduced by Goodfellow et al.[2], are a powerful class of generative models that learn to approximate complex data distributions through a two-player minimax game between a generator and a discriminator. The adversarial nature of this setup drives the generator to produce highly realistic samples so that the discriminator struggles to identify them as synthetic. Since their introduction, several theoretical improvements have been proposed to stabilize training and improve convergence, including Wasserstein GANs [3]. Due to their ability to generate high-fidelity synthetic samples, GANs have been increasingly applied to the healthcare domain, including Electronic Health Records (EHR) and longitudinal data, where they are considered promising for tasks such as privacy-preserving data sharing and data augmentation. Recent reviews have surveyed a wide range of GAN-based models adapted to clinical data highlighting their potential and outlining common evaluation metrics and benchmark datasets [4]. Given the characteristics of our use case, we excluded models that do not handle longitudinal EHR data or are specifically designed for generating continuous-valued time-series

#### *2 T-GPT*

T-GPT adapts autoregressive Transformer modeling to patient-level tabular clinical/registry data by serializing each row into a sequence of field tokens and training with a next-token objective. This leverages self-attention to capture cross-feature dependencies without recurrence, following the original Transformer formulation [5]. In practice, the row is linearized with special separators and field identifiers so that the model conditions on previously generated fields; this “table-to-sequence” strategy is widely used in Transformer tabular generators such as REaLTabFormer (GPT-style decoding for parent tables, then Seq2Seq for related tables) and GReaT (LLM fine-tuned to emit rows), which also describe practical tokenization choices for categorical and numeric attributes [6]. Within the broader tabular-Transformer literature, several designs explain why attention is effective for tables and inform T-GPT’s column-wise conditioning: TabTransformer learns contextual embeddings for categorical columns via stacked self-attention [7]; FT-Transformer shows that a simple feature-tokenization plus attention baseline competes strongly across diverse tabular tasks [8]; and SAINT introduces row- and column-attention with contrastive pretraining, reinforcing the value of attention over both samples and features [9]. For scenarios involving evolving schemas or multiple tables, TransTab demonstrates that converting cells and column descriptions into sequences enables transfer across tables, an approach compatible with T-GPT’s serialization and conditional generation (e.g., prefixing demographic or cohort constraints before sampling) [10].

### SUPPLEMENTARY_FILE_4 (SF4) – Data pre-processing

From 79,001 patients available at extraction, application of these criteria yielded an analysis cohort of 4,878 individuals, of whom 914 initiated an early-intensive treatment (EIT) and 3,964 followed an escalation strategy (ESC). For treatment grouping, EIT included natalizumab, alemtuzumab, ocrelizumab, cladribine, fingolimod, or mitoxantrone as initial DMTs, whereas ESC included interferon-beta products, glatiramer acetate, teriflunomide, or dimethyl fumarate with subsequent escalation to a high-efficacy agent after at least one year. Outcomes and time origin were harmonized as follows: baseline was the date of first DMT; confirmed disability accrual (CDA) was defined as a 6-month confirmed EDSS increase with thresholds dependent on baseline EDSS (≥1.5 if baseline=0; ≥1.0 if baseline >1.0 and <5.5; ≥0.5 if baseline >6.0), with the CDA date assigned at the first EDSS showing the increase; the primary endpoint, progression independent of relapse activity (PIRA), was defined as a CDA event occurring >90 days after and >30 days before a relapse. Variables used in downstream models comprised sex, age at first DMT, EDSS at first DMT, time from onset to first DMT, number of relapses in the two years before treatment (0/1/≥2), onset type (monofocal/multifocal), T2-lesion categories at the pre-treatment MRI (0, 1–2, 3–8, ≥9), presence/absence of gadolinium-enhancing T1 lesion, and treatment strategy (ESC as reference). Time-to-event analyses employed Cox proportional hazards models to estimate risk of PIRA in both real and synthetic cohorts of 4,878 subjects.

### SUPPLEMENTARY_FILE_5 (SF5) – Training strategies and implementation details

We implemented the two generative approaches: a Generative Adversarial Network (GAN) and a Tabular Generative Pretrained Transformer (T-GPT) mentioned above, both trained on the sub-cohort (n=1,666) extracted from the Italian MS Register, and used them to synthesize a cohort matching the analysis size (n=4,878). The dataset was split into training (80%) and testing (20%).

The transformer-based model was a GPT-2 architecture fine-tuned for tabular synthesis, while the GAN served as a complementary generator to benchmark performance across families of models. Model development and selection were guided by the SAFE validation framework. Feature encoding followed the native tabular representation used for clinical and MRI variables in RISM, with categorical fields modeled as discrete features and continuous fields retained as scalars; temporal quantities (e.g., time to first DMT start and time to first PIRA) were included as scalar features rather than sequences.

Both model hyperparameters were chosen based on training stability and a qualitative evaluation of the generated samples. Models training parameters and settings are reported below in Tables 4 and 5.

Table 4: T-GPT model and training parameters used in this study

| **Parameter** | **Value** | **Description** |
|---|---|---|
| Batch size | 32 | Training batch size |
| Max epochs | 200 | Maximum number of training epochs |
| Learning rate | 1e-4 | Learning rate |
| Embedding size | 256 | Size of code embeddings |
| Transformer layers | 8 | Number of Transformer layers |
| Attention heads | 8 | Number of attention heads |
| Layer norm epsilon | 1e-5 | A small constant used for stabilizing layer normalization. |

Table 5: GAN model and training parameters used in this study

| **Parameter** | **Value** | **Description** |
|---|---|---|
| Batch size | 64 | Batch size used during GAN training. |
| Iterations | 30000 | Number of training iterations for GAN training. |
| Generator hidden size | 256 | Hidden dimension size in the generator network. |
| Generator attention size | 128 | Dimension of attention layer in the generator. |
| Critic hidden size | 128 | Hidden dimension size in the critic (discriminator). |
| Noise size | 256 | Size of noise vector for generator. |

| **Parameter** | **Value** | **Description** |
|---|---|---|
| Generator iterations | 2 | Number of generator updates per training step. |
| Generator learning rate | 5e-5 | Learning rate for the generator optimizer. |
| Critic iterations | 1 | Number of critic (discriminator) updates per training step. |
| Critic learning rate | 5e-5 | Learning rate for the critic optimizer. |
| Beta0, Beta1 | 0.5, 0.9 | $\beta 0$ and $\beta 1$ parameters for the Adam optimizer. |
| Lambda | 10 | Gradient penalty coefficient |
| Decay rate | 0.5 | Learning rate decay factor. |
| Decay step | 10000 | Step interval for applying learning rate decay. |
| Base GRU epochs | 200 | Number of training epochs for the base GRU model. |
| Base GRU learning rate | 1e-3 | Learning rate for the base GRU model. |

**SUPPLEMENTARY_FILE_6 (SF6) – Development of SAFE**

Assessing the quality of the generated data is a notoriously challenging task. A Synthetic vAlidation FramEwork (SAFE) powered by Train was implemented to assess the models and quality of the synthetic patients with respect to its fidelity, correlation, utility, and privacy [11; 12; 13; 14; ?SafeREF?]. SAFE handles both continuous and discrete, constant or time-varying variables. SAFE integrates and extends methods from existing frameworks for synthetic data evaluation, aiming to address key limitations in their application to longitudinal and domain-specific clinical contexts. Several established tools support synthetic data evaluation, including the Synthetic Data Vault (SDV) [15], an open-source library that provides generators and metrics for single-table, multi-table, and time-series data. While SDV includes useful fidelity and privacy indicators, it lacks clinical-specific validation components. Similarly, SynthEval [16] offers a suite of utility, privacy, and fairness metrics for tabular data, but does not support longitudinal structures or clinical relevance assessments. The YData SDK [17] provides tools for data profiling and synthetic data generation, along with a Synthetic Data Quality Report that summarizes privacy, utility, and fidelity scores; however, it does not include methods tailored to healthcare applications. SAFE builds on and integrates capabilities inspired by these tools, while introducing domain-aware validation strategies designed for use in clinical research. In addition to standard statistical indicators, SAFE incorporates validations based on clinical endpoints, such as survival analysis using Kaplan–Meier curves and log-rank tests, rule-based consistency checks, and outcome comparisons between real and synthetic cohorts. This enables rigorous assessment of both temporal fidelity and clinical plausibility, which are essential in healthcare contexts. To ensure a comprehensive and extensible evaluation pipeline, SAFE leverages several state-of-the-art statistical and machine learning libraries. These include scikit-learn [18], which provides a comprehensive set of metrics for machine learning evaluation, lifelines [19] for survival analysis, scipy.stats [20] for statistical hypothesis testing, statsmodels [21] for time-series modeling and causality analysis, and pandas [22] for data manipulation and correlation assessment. By combining these general-purpose tools with clinical-specific validations, SAFE offers a robust and versatile framework for evaluating synthetic healthcare data.

*1 Fidelity metric*

SAFE calculates a fidelity metric ranging from 0 to 1, where values closer to 1 indicate higher fidelity. First, the dimensional probability test compares all binary variable probabilities over all visits of synthetic and real data. Next, the probabilities, per record and per visit, of unigrams (single events) and bigrams (pairs of events) are compared, considering both pairs within the same visit and between consecutive visits. The values of the coefficient of determination $R^2$ are calculated and the $max(0, R^2)$ are averaged for the calculation of the *fidelity_time_discrete_prob_all* metric in the case of the dimensional probability test and the *fidelity_time_discrete_prob_rec_vis* metric in the case of probabilities per record and per visit. In addition, the correlation coefficient (CC) and root mean squared error (RMSE) are also computed to support the analysis. For continuous variables, the Max Mean Discrepancy (MMD), a kernel-based statistical measure used to compare two data distributions, has been employed to verify that real and synthetic data originate from the same distribution. The value of the Max Mean Discrepancy was used to construct the metric with values from 0 to 1 *fidelity_time_continuous_MMD*= $1/c^d$, with $d$ representing the discrepancy between the distributions and $c$ set to 1.2 by verifying with test data and random data that, when they come from the same distribution, it gives values close to one, and when they are far apart, it tends to zero.

For time-constant discrete variables, the TVD (Total Variation Distance) is computed and 1-TVD becomes the component of the fidelity metric *fidelity_constant_discrete*, which gives a similarity score the synthetic data and real data distributions: values closer to 1 imply that the distribution of the synthetic data is highly similar to that of the real data, while values closer to 0 imply a greater difference between the distributions. For the distributions of time-constant continuous variables and patient lengths (as number of visits), the Kolmogorov Smirnov (KS) test is applied, the components of the metrics *fidelity_lengths* and *fidelity_constant_continuous* consist of 1- KS statistic. Another component of the fidelity metric was developed to analyze mean values across visits for continuous variables and frequencies for discrete variables, which includes SMAPE (Symmetric mean absolute percentage error), Granger causality test and correlation. SMAPE is transformed into a metric ranging from zero to one by computing the mean SMAPE across all variables and applying the transformation $max(0, 1 - mean(SMAPE))$.

The correlation is converted to a metric *fidelity_across_visits,* with values in [0, 1], by averaging $max(0, x)$ where $x$ represents the individual correlation results for each variable. The Granger causality test is performed, saving for each variable the minimum p-value across all tests and lags, and the average complement of p-values is calculated, giving a score where higher values indicate stronger causality. The three values are then averaged into a single metric. This is done for original discrete variables, i.e. binary variables active only at the time of detection first forward and then backward, as well as for the switch variables, where only changes in categorical variables are considered (equal to 1 in status changes eg. from negative to positive). In addition, the assessment of the fidelity of the distributions per visit is conducted with the Total Variation Distance for discrete data or the Kolmogorov-Smirnov test for continuous data. Again, the averages of 1- TVD and 1- KS statistic, calculated per variable and per visit, are used for the metrics *fidelity_per_visit_discrete* and *fidelity_per_visit_continuous*.

A final fidelity analysis is performed by creating Uniform Manifold Approximation and Projection (UMAP) embeddings and analysing the mapping of real and synthetic data and the positions over time of patients in the new space. For each visit, UMAP embeddings of real and synthetic are calculated for each visit using dice metric for categorical variables (already converted to one-hot encoding format) and Euclidean metric on standardised continuous variables, and the two fits (numerical and categorical) are summed. Afterwards, for each synthetic patient, the closest real patient at the first visit is found, considering the filled variables, i.e. starting with the same tumour characteristics. Distances in the UMAP spaces from the patient identified as the closest are plotted over time. The UMAP embeddings in the visits are plotted in 2D and 3D for graphical analysis. In addition, outlier patient IDs, found as those having at least one distance in time that is an outlier, based on upper bound IQR of distances, are identified and saved. The various components of fidelity are averaged to obtain fidelity metrics for constant variables, patient lengths, time-varying continuous variables, and time-varying discrete variables. These are subsequently weighted according to the number of variables in each category to derive a single final score.

*2 Correlation analysis*

The relationships between the columns of the dataset were evaluated by building correlation matrices. Since the clinical and demographic features had different data types (i.e. continuous and categorical), we used different methods to calculate the correlation metric. We calculated the correlation of continuous-continuous features using Pearson's correlation. For continuous-categorical features, we used the Correlation Ratio and for categorical-categorical features we performed the Uncertainty Coefficient proposed by Theil's U. We created two correlation matrices for real and synthetic datasets, then we calculated the difference between those matrices. Metric was calculated as the average of all values in the difference matrix.

*3 Privacy metric*

The privacy metric, with values between 0 and 1, results from a membership inference attack aimed at inferring whether a real record was used in the training dataset. Having labeled the training and test data as positive and negative, the attack is carried out by calculating the distances of the synthetic dataset from both. The distances are calculated with Hamming distance for the categorical variables, and the Euclidean distance for the continuous variables, after normalization by min-max scaling, and combined as a weighted sum, with weights proportional to the number of variables. Then data with distances lower than the median are classified as positive ("in training"), otherwise as negative ("not in training") . Accuracy, precision, and recall are then calculated by comparing these predicted labels to the ground truth. A perfectly private synthetic dataset would yield 50% for all three metrics, indicating random guessing. The final privacy score is derived as $1 - 2 \cdot \text{mean}(|\text{metric} - 0.5|)$, averaged over accuracy, precision, and recall, where a score close to 1 indicates stronger privacy. Furthermore, a visual privacy assessment is done with the empirical cumulative distribution of the minimum distances between real-real, synthetic-synthetic, and synthetic-real. For a quantitative assessment, the averages of the minimum distances in the 3 cases are printed.

*4 Clinical validation*

A metric was developed and implemented in the SAFE library to account for clinical validation: Clinical Synthetic Fidelity (CSF). It summarizes multiple tests that compare, variable by variable, the distributions observed in the real cohort against those in the synthetic cohort, yielding a single score (optimal ≥90).

### SUPPLEMENTARY_FILE_7 (SF7) – Experimental results settings

Evaluation of our generative approach was carried out by running a battery of experiments with the SAFE library to assess three aspects: (i) fidelity of the synthetic cohort to real-world clinical distributions, (ii) structure preservation via correlation analysis, and (iii) privacy protection. Clinical utility was then tested by reproducing the comparative-effectiveness analysis with identical Cox models in real and synthetic data.

#### 1 Fidelity

Fidelity was quantified with the Clinical Synthetic Fidelity (CSF) metric implemented in SAFE. In our study, CSF was 97% (optimal ≥90%, as cited above), indicating excellent alignment of the synthetic cohort with the empirical distributions of the registry data. In Figure 1, the distributions of variables are shown, including age, sex, time to first disease-modifying-therapy (DMT) start, PIRA, time to PIRA, T1 Gadolinium (GD) lesions at baseline, T2 lesions at baseline, baseline EDSS, treatment strategies and number of relapse prior to DMT.

Figure 1: Distributions of synthetic vs real data variables. Blue illustrates the real data, while red illustrates synthetic data.

Abbreviations: DMT, PIRA, GD, EDSS, ESC, EIT (see Table 1 in Supplementary file for more information)

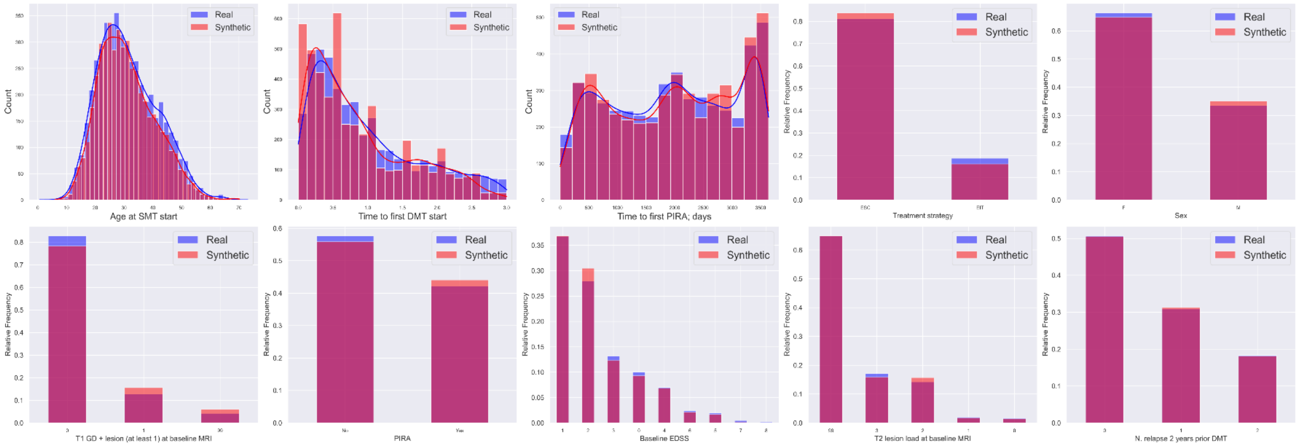


#### 2 Correlation

To verify that relationships among variables were preserved, we computed correlation matrices separately in real and synthetic data, choosing the appropriate association measure by type: Pearson for continuous–continuous, correlation ratio for continuous–categorical, and Theil’s U (uncertainty coefficient) for categorical–categorical pairs. We then formed a difference matrix (real minus synthetic) and summarized preservation as the average value in this difference matrix. In our results, both the direction and strength of associations were highly consistent across datasets and the differences were close to zero. Figure 2 shows the two correlation heatmaps and the difference heatmap.

Figure 2: Correlation heatmaps and difference heatmap plots.

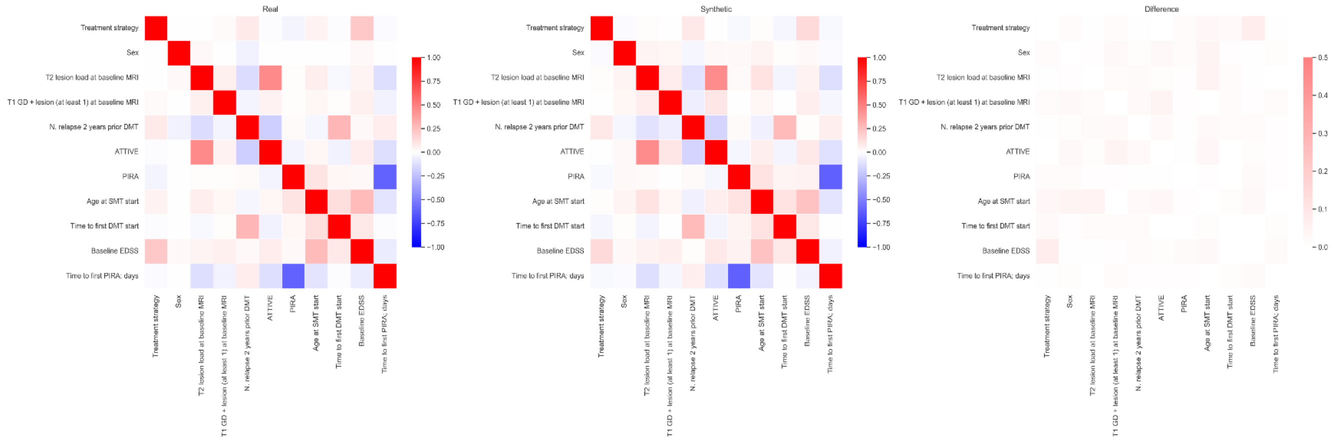

*3 Privacy*

Privacy was evaluated with the Nearest Neighbor Distance Ratio (NNDR), with values in the 0.60–0.85 range considered optimal. The synthetic cohort achieved NNDR = 0.61, consistent with privacy preservation and absence of identifiable leakage.

*4 Clinical validation and utility*

Clinical validation used the Clinical Synthetic Fidelity (CSF) from SAFE as a primary, task-oriented metric summarizing concordance between real and synthetic clinical distributions (97% in this study). To further test clinical utility, we fit identical Cox proportional-hazards models for time to first PIRA in both cohorts, including covariates: sex, age and EDSS at first DMT, time from onset to first DMT, relapses in the prior two years (0/1/≥2), onset type (mono- vs multifocal), T2 lesion categories (0, 1–2, 3–8, ≥9), presence of GD-enhancing T1 lesion at baseline MRI, and treatment strategy (ESC reference). Proportional-hazards assumptions were checked with log-time interactions and Schoenfeld residuals; results were reported as HRs with 95% CIs.

The comparative-effectiveness signal was consistent across datasets: early intensive therapy (EIT) vs escalation (ESC) showed HR = 0.88 (95% CI 0.71–1.08, p = 0.22) in real data and HR = 0.86 (95% CI 0.76–0.98, p = 0.02) in synthetic data. Age at DMT start increased PIRA risk in both datasets (HR≈1.02 per year, p<0.001), while in synthetic data a longer time to first DMT was also associated with higher risk (HR = 1.10, 95% CI 1.04–1.17, p<0.001). MRI-derived factors retained interpretable effects in synthetic data (≥9 T2 lesions: HR = 1.88, 95% CI 1.20–2.96; GD-enhancing (GD +) T1 lesion: HR = 0.82, 95% CI 0.73–0.91), mirroring clinical patterns even when not significant in the real cohort. Full HRs with 95% CIs as well as reference levels in categorical variables included in the model are summarized in Table 6 (real cohort) and Table 7 (synthetic cohort); an overview is also shown in the forest plots attached in the main manuscript.

Table 6: Cox model HRs and CIs (real cohort)

| Feature | Nº of patients | HR | p-value | Lower CI | Upper CI |
|---|---|---|---|---|---|
| Age at DMT start | 4878 | 1,0181 | 5,9265e-06 | 1,0102 | 1,0261 |
| Sex: Female | 3241 | Reference | | | |
| Sex: Male | 1637 | 0,9672 | 0,6872 | 0,8223 | 1,1376 |

| | | | | | |
|---|---|---|---|---|---|
| Type of Disease Onset: Monofocal | 4045 | Reference | | | |
| Type of Disease Onset: Multifocal | 626 | 1,1213 | 0,2802 | 0,9109 | 1,3802 |
| Numb. of Relapse before DMT start: 0 | 2476 | Reference | | | |
| Numb. of Relapse before DMT start: 1 | 1507 | 0,9213 | 0,3690 | 0,7705 | 1,1017 |
| Numb. of Relapse before DMT start: > 2 | 895 | 1,1195 | 0,2800 | 0,9122 | 1,3741 |
| EDSS at DMT start | 4878 | 0,9720 | 0,3959 | 0,9105 | 1,0378 |
| Time to first DMT start | 4878 | 1,0995 | 0,0744 | 0,9907 | 1,2202 |
| Treatment: ESC | 3964 | Reference | | | |
| Treatment: EIT | 914 | 0,8760 | 0,2225 | 0,7082 | 1,0836 |
| T1 GD + lesions: No | 2634 | Reference | | | |
| T1 GD + lesions: Yes | 877 | 0,9489 | 0,5443 | 0,8009 | 1,1242 |
| T2 Brain lesions load: 0 | 80 | Reference | | | |
| T2 Brain lesions load: 1-2 | 101 | 1,2661 | 0,3413 | 0,7788 | 2,0583 |
| T2 Brain lesions load: 3-8 | 695 | 1,2421 | 0,2804 | 0,8379 | 1,8414 |
| T2 Brain lesions load: >= 9 | 836 | 1,2553 | 0,2536 | 0,8497 | 1,8545 |

Table 7: Cox model HRs and CIs (synthetic cohort)

| Feature | Nº of patients | HR | p-value | Lower CI | Upper CI |
|---|---|---|---|---|---|
| Age at DMT start | 4878 | 1,0173 | 6,2011e-14 | 1,0128 | 1,0219 |
| Sex: Female | 3339 | Reference | | | |
| Sex: Male | 1539 | 1,0145 | 0,7617 | 0,9244 | 1,1134 |
| Type of Disease Onset: Monofocal | 4372 | Reference | - | - | - |

| | | | | | |
|---|---|---|---|---|---|
| Type of Disease Onset: Multifocal | 506 | 0,9935 | 0,9285 | 0,8622 | 1,1448 |
| Numb. of Relapse before DMT start: 0 | 2521 | Reference | - | - | - |
| Numb. of Relapse before DMT start: 1 | 1500 | 0,8412 | 0,0009 | 0,7596 | 0,9316 |
| Numb. of Relapse before DMT start: > 2 | 857 | 0,9700 | 0,6284 | 0,8575 | 1,0973 |
| EDSS at DMT start | 4878 | 1,0169 | 0,3655 | 0,9806 | 1,0545 |
| Time to first DMT start | 4878 | 1,1040 | 0,0012 | 1,0399 | 1,1720 |
| Treatment: ESC | 4130 | Reference | - | - | - |
| Treatment: EIT | 748 | 0,8603 | 0,0223 | 0,7562 | 0,9788 |
| T1 GD + lesions: No | 3816 | Reference | - | - | - |
| T1 GD + lesions: Yes | 1062 | 0,8160 | 0,0003 | 0,7307 | 0,9112 |
| T2 Brain lesions load: 0 | 71 | Reference | - | - | - |
| T2 Brain lesions load: 1-2 | 115 | 1,4935 | 0,1458 | 0,8698 | 2,5643 |
| T2 Brain lesions load: 3-8 | 1875 | 1,5626 | 0,0551 | 0,9902 | 2,4658 |
| T2 Brain lesions load: >= 9 | 2817 | 1,8838 | 0,0062 | 1,1969 | 2,9650 |
